# Efficacy of N-163 strain of *Aureobasidium pullulans*-produced beta-glucan in improving muscle strength and function in patients with Duchenne muscular dystrophy; Results of a 6-month non-randomised open-label linear clinical trial

**DOI:** 10.1101/2023.04.29.23289260

**Authors:** Kadalraja Raghavan, Thanasekar Sivakumar, Sudhakar S Bharatidasan, Subramaniam Srinivasan, Vidyasagar Devaprasad Dedeepiya, Nobunao Ikewaki, Rajappa Senthilkumar, Senthilkumar Preethy, Samuel JK Abraham

**Author notes:** **Corresponding author information:** Dr. Samuel JK Abraham, II Department of Surgery & CACR, University of Yamanashi, Faculty of Medicine, Address for correspondence: 3-8, Wakamatsu, Kofu, 400-0866, Yamanashi, Japan, Email id-; Alternate email id. **Declarations:**. ***Ethics approval and consent to participate*** The study was registered in Clinical trials registry of India, CTRI/2022/05/042905 and approved by the ethics committee of Saravana Multispeciality Hospital-Institutional Ethics Committee, India. ***Consent for publication*** Not applicable. ***Availability of data and material*** All data generated or analysed during this study are included in the article itself. ***Funding*** No external funding was received for the study. ***Competing interests*** Author Samuel Abraham is a shareholder in GN Corporation, Japan which holds shares of Sophy Inc., Japan., the manufacturers of novel beta glucans using different strains of Aureobasidium pullulans; a board member in both the companies and also an applicant to several patents of relevance to these beta glucans. ***Authors’ contributions*** KR, NI and SA. contributed to conception and design of the study. KR and RS helped in data collection and analysis. SA and SP drafted the manuscript. VD, SS and SB performed critical revision of the manuscript. All authors read and approved the final manuscript.

## Abstract

Duchenne muscular dystrophy (DMD) is a debilitating genetic disease that causes gradual muscle weakening and early mortality. After a 45-day clinical pilot study and a pre-clinical study in mdx mice demonstrating safety and efficacy, and anti-fibrotic and anti-inflammatory effects, respectively, of an *Aureobasidium pullulans* N-163 strain-produced B-1,3–1,6-glucan. We conducted this linear six-month clinical study to assess its efficacy. Twelve ambulatory and 12 non-ambulatory individuals with DMD were included; all received the N-163 strain of A.pullulans produed beta-glucan dietary supplement orally in addition to the standard treatment regimen, which included steroids. The Medical Research Council muscle score improved in 11 patients in the ambulatory group and in 8 patients in the non-ambulatory group. The six-minute walk test distance improved in nine patients, with a 29.5-meter average improvement. The North Star Ambulatory Assessment improved by 1 unit in three patients. This safe beta-glucan food supplement improved muscle function within 6 months. A comprehensive, multi-centric clinical research should be conducted for unravelling its potential as a disease-modifying drug adjuvant in DMD.

## Introduction

Duchenne muscular dystrophy (DMD) is a lethal X-linked recessive disease, whose prevalence varies between 1 in 3600 and 1 in 9300 male infants globally.^1^ Loss-of-function mutations in the DMD gene prevent the production of functional dystrophin protein in patients with DMD, which causes progressive skeletal muscle weakening and degeneration, heart dysfunction, respiratory failure, and ultimately premature death.^1,2^ The two treatment approaches for DMD are as follows: the first is dystrophin-targeted therapy, and the alternative is to focus on the pathological alterations that occur downstream. Gene, cell, and protein replacement therapies are examples of treatments that target dystrophin. However, dystrophin-targeted therapy can only slow the course of the disease and cannot restore the function of defective muscle tissues. Second, owing to the body’s widespread distribution of muscle tissue, dystrophin-targeted medicines have difficulty reaching all muscle tissues.^3^ Targeting secondary downstream pathological mechanisms is another therapeutic approach. Multiple pathogenic processes, such as fibrosis, inflammation, loss of calcium homeostasis, oxidative stress, ischaemia, and muscular atrophy, are triggered by downstream pathological mechanisms.^3^ Different approaches that target fibrosis, inflammation, ischaemia, etc., are available, although a wholesome approach to DMD that aims both at dystrophin replacement as well as addressing downstream pathological mechanisms is necessary.

Previously, we have reported on a 1-3,1-6 beta-glucan from the N-163 strain of the black yeast *Aureobasidium pullulans* that helped decrease interleukin (IL)-6, tumour growth factor (TGF)-β, and IL-13 and increase dystrophin levels along with improvement of muscle strength in patients with DMD in a clinical study performed in 28 patients with DMD for a duration of 45 days.^4^ In the same study, we reported the beneficial constitution of the gut microbiome with an increase in butyrate-producing species such as *Roseburia* and *Faecalibacterium prausnitzii*, apart from a decrease in harmful bacteria associated with inflammation such as enterobacteria and *Alistipes*.^5^ We also conducted a pre-clinical study of this N-163 beta-glucan in an mdx mice model, which revealed a significant decrease in inflammation score and fibrosis as well as levels of plasma alanine transaminase, aspartate aminotransferase, lactate dehydrogenase, IL-13, and haptoglobin; increased anti-inflammatory TGF-β levels; and balanced regulation of the quantity of centrally nucleated fibres indicative of muscle regeneration.^6^ In a pre-clinical study of mdx mice, we also reported muscle regeneration and maturation by MYH3 and CD44 expression.^7^ In the present study, we evaluated the effects of N-163 beta-glucan for 6 months in patients with DMD.

## Materials and methods

This trial was an investigator-initiated, single-centre, randomised, open-label, prospective, linear, single-arm clinical study of patients with DMD. This study was conducted over a period of 180 days.

The intervention was consumption of one sachet of N-163 beta-glucan (Commercial name: Neu-REFIX) once daily along with conventional treatment comprising routine physiotherapy for joint mobility along with medications, viz., T. calcium and vitamin D 1000 with or without deflazacort (steroid) 6–24 mg.

### Inclusion criteria

This study included male patients with a molecular diagnosis of DMD aged 3–30 years who were willing to participate in the study with written informed consent.

### Exclusion criteria

Patients with a previous (within the past 1 month) or concomitant participation in any other therapeutic trial; a known or suspected malignancy; any other chronic disease, or clinically relevant limitation of renal, liver, or heart function according to the discretion of the investigator were excluded from the study.

### Investigations

A background survey was conducted to obtain data on sex, date of birth, age, habits, current medical history, medication, treatment, allergies (to drugs and food), regular use of food for specified health uses, functional foods, health foods, intake of foods rich in beta-glucan and foods containing beta-glucan, and intake of immunity-boosting foods. Medical history and physical measurements were taken, including height, weight, body mass index, and temperature. The following tests were performed at baseline and at the end of the study (6 months).

1. Physiological examination included assessing the systolic blood pressure, diastolic blood pressure, and pulse rate.
2. Electrocardiogram (ECG).
3. The Medical Research Council (MRC) Scale was used to measure muscle strength.
4. six-minute walk test (6MWT) and North Star Ambulatory Assessment (NSAA)

The participants were contacted every week for drug compliance and the recording of adverse effects, if any.

At 3 and 6 months, a physiological examination was performed to assess systolic blood pressure, diastolic blood pressure, and pulse rate.

### Statistical analysis

Microsoft Office Excel statistics package and the software Origin 2021b were used for statistical analysis. Statistical significance was set at p<0.05, and multiple comparisons or multiple outcomes were not corrected.

## Results

Altogether, 27 patients were screened, of whom 24 were included in the study. The mean ± standard deviation of age for the total study population was 11.33 ± 4.57 years (range, 5–26 years).

No adverse events were observed. No clinically significant changes from baseline data were observed on physical examination or in vital signs—temperature, blood pressure, oxygen saturation, pulse rate, or ECG (data not shown).

The mean NSAA score at pre-intervention (21.41 ± 11.81) and post-intervention (21.58 ± 11.87) did not indicate any significant change. The NSAA score improved by 1 unit in three patients and decreased by 1 unit in one patient. In the remaining patients, no change was identified (Figure 1). In the ambulatory group (n=12), the MRC score improved in 11 patients, with an average improvement of 7.09%, while it increased by 9.27% in 8 of the 12 non-ambulatory patients and decreased in 2 cases (Figure 2). The 6MWT distance improved in 9 patients, with a 29.5-metre average improvement (Figure 3).

**Figure 1:**
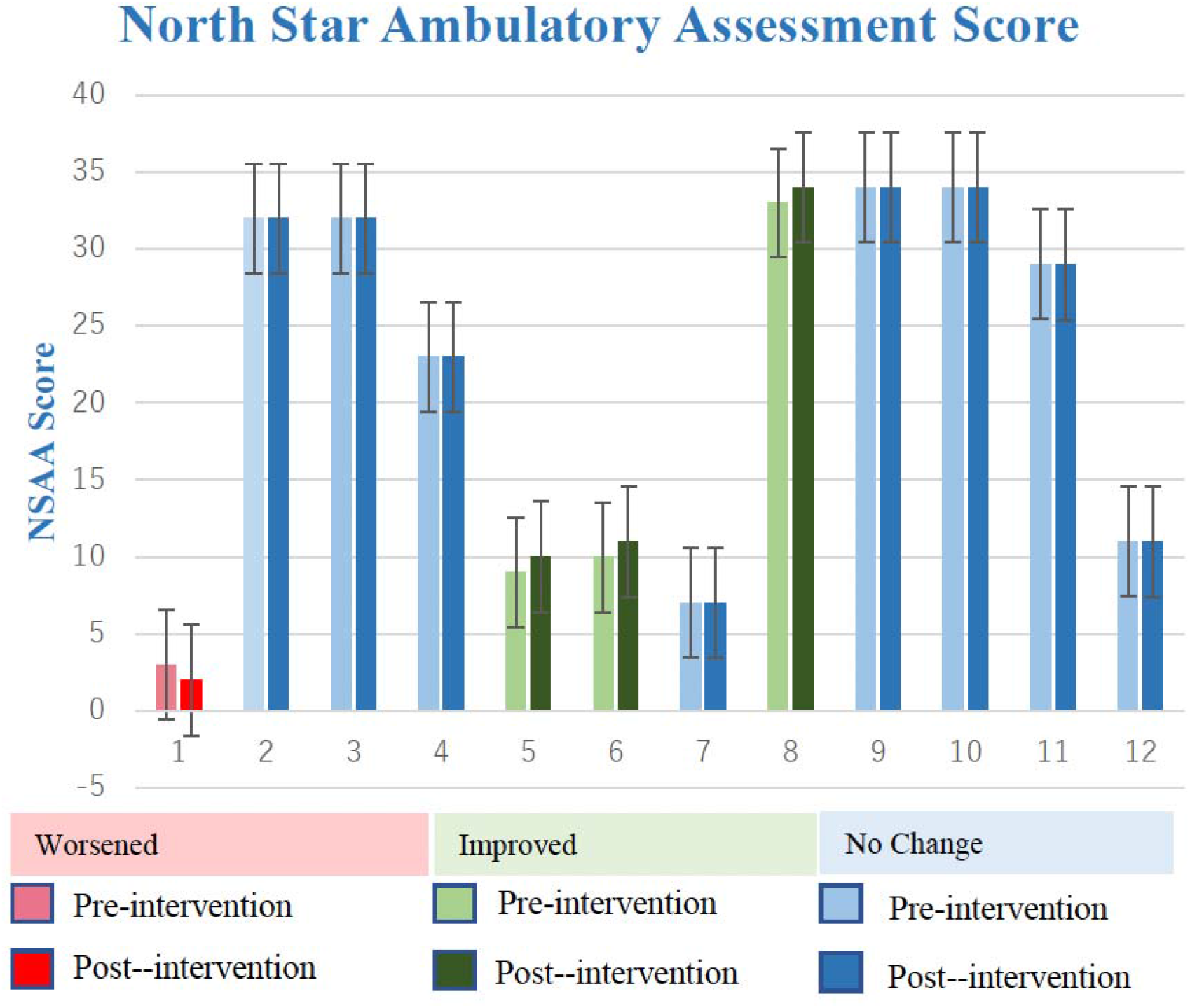
North Star Ambulatory Assessment Score pre- and post-intervention of N-163 beta-glucan in patients with Duchenne muscular dystrophy.

**Figure 2:**
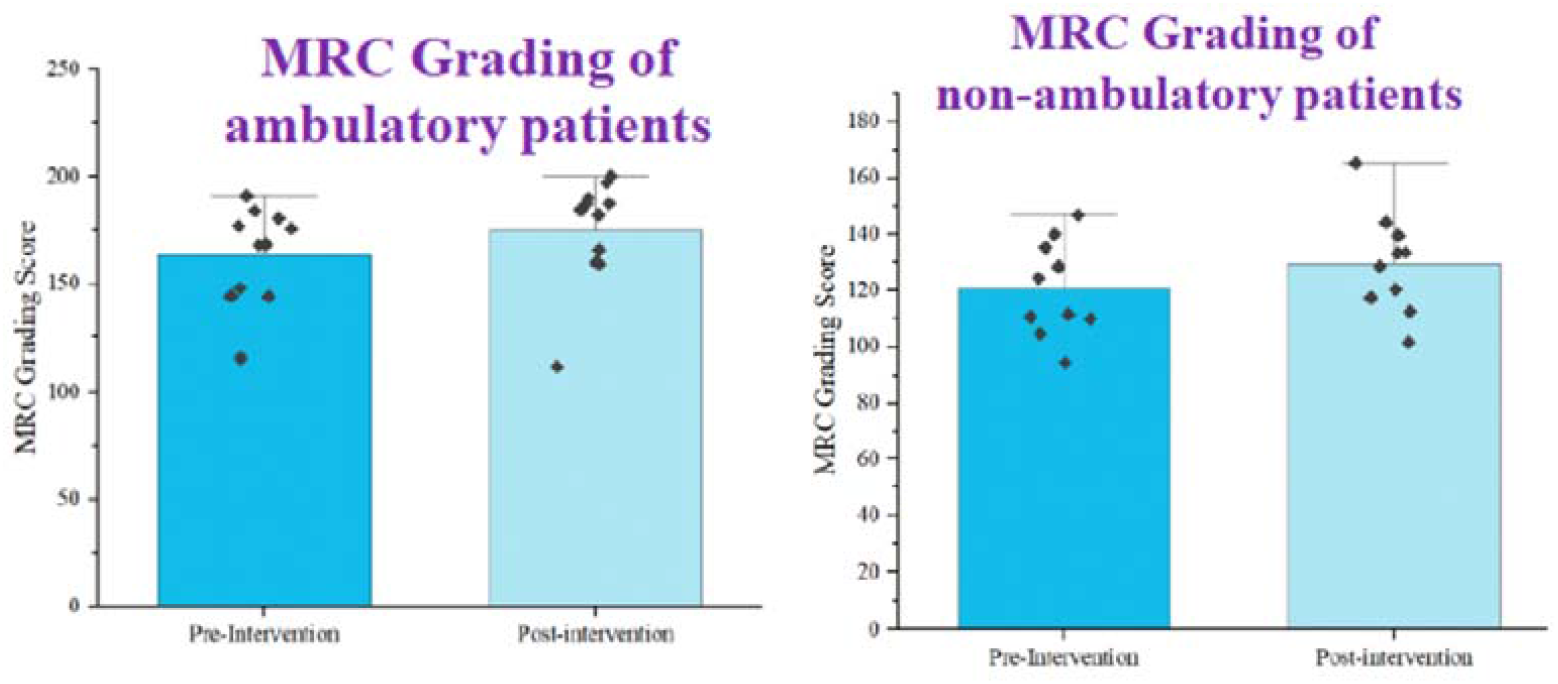
Medical Research Council grading pre- and post-intervention of N-163 beta-glucan in patients with Duchenne muscular dystrophy.

**Figure 3:**
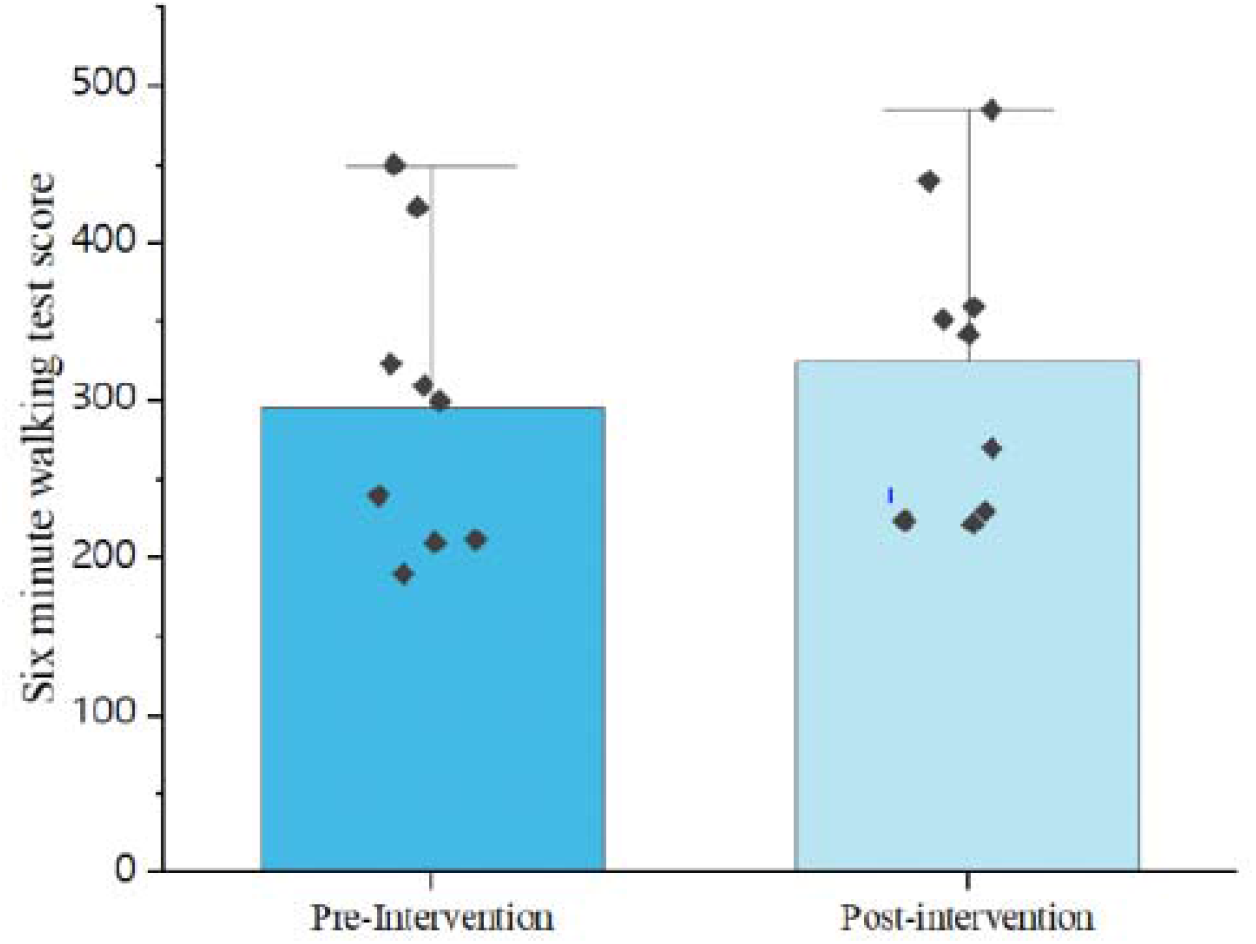
Six-minute walk test score pre- and post-intervention of N-163 beta-glucan in patients with Duchenne muscular dystrophy.

## Discussion

The goal of DMD treatment is to correct genetic defects and restore muscle function apart from re-establishing normal levels of dystrophin.^1-3^ Despite some approaches being in clinical trials, gene correction therapies such as exon-skipping therapies are available only for a select group of patients with specific exon deletions. Although supportive treatments aim to halt or delay the disease progression, they do not significantly contribute to muscle regeneration and the restoration of muscle function, apart from the associated side effects. Even in long-term studies, only in terms of delaying the disease progression, positive outcome was reported without any improvement of muscle strength or function after the intervention compared to the baseline data in the treatment arm^8^. One of the reasons for the lack of adequate muscle regeneration is inflammation.^1-3^ Since we have earlier proven the decrease in inflammation through biomarker levels in clinical and pre-clinical studies,^4-6^ restoration of gut microbiome^6^, and muscle regeneration after consumption of N-163 strain of *Aureobasidium pullulans* produced beta-glucan evident from increased MYH3 expression and proper muscle maturation by increased CD44 hyaluronan expression in an mdx mice model,^7^ which could be attributed to the clinical improvement in muscle strength and function in the present study.

In our present study of only six months, the 6MWT distance improved in 9 patients, with a 29.5-metre average improvement. Although the NSAA scores did not indicate any significant differences pre- and post-intervention, the improvement in MRC grading in 91.6% of the participants, apart from improvement in the 6MWT indicates the significance of this N-163-produced beta-glucan approach in terms of muscle function improvement, which is not commonly observed with conventional or even other recent therapeutic approaches.

The limitations of this study are its smaller sample size and heterogeneity of the standard treatment regimens followed and a duration of six-months. Nevertheless, the safety and muscle function improvement efficacy of this N-163-produced beta-glucan food supplement without adverse reactions can be considered an indispensable addition to disease-modifying treatments for DMD and other dystrophies, such as limb-girdle muscular dystrophy.

## Conclusion

This safe N-163 strain of *Aureobasidium pullulans* produced beta-glucan food supplement is a promising disease-modifying pharmacological adjuvant in DMD, as it improved MRC in 91.6% of the patients, with modest improvements in 6MWT and NSAA score after 6 months. In the future, a comprehensive, multi-centric clinical study should be conducted to unravel the potentials of this orally consumable, safe, and allergen-free food supplement as a drug adjuvant in delaying the progression of DMD and other dystrophies.

## Data Availability

All data produced in the present work are contained in the manuscript

## Acknowledgements

The authors thank

1. The Government of Japan and the Prefectural Government of Yamanashi for a special loan and M/s Yamanashi Chuo Bank for processing the transactions.
2. Ms. Sunitha, Mr. Vincent and the staff of JAICARE and Sarvee Integra, Dr.
3. Ragaroobine, Mr. Rajmohan from Nichi-In Centre for Regenerative Medicine (NCRM) for their assistance during the clinical study and data collection of the manuscript.
4. Fr. Francis Xavier, Rector, Loyola College, Chennai, Fr. Vargheesh Antony and Fr.
5. Marianathan of JAICARE for their support during the clinical study.
6. Ms. Misa Takamoto, Mr. Masato Onaka, Mr. Yasushi Onaka of Sophy Inc, Kochi, Japan for necessary technical support.
7. Ms. Yoshiko Amikura and staff of GN Corporation, Japan for their liaison assistance with the conduct of the study.
8. Loyola-ICAM College of Engineering and Technology (LICET) for their support to our research work.

## Abbreviations

DMD: Duchenne muscular dystrophy
6MWT: Six-minute walk test
NSAA: North Star Ambulatory Assessment
MRC: Medical Research Council

